# Population attributable fraction for continuously distributed exposures

**DOI:** 10.1101/2020.10.15.20212886

**Authors:** John Ferguson, Fabrizio Maturo, Salim Yusuf, Martin O’Donnell

## Abstract

When estimating population attributable fractions (PAF), it is common to partition a naturally continuous exposure into a categorical risk factor. While prior risk factor categorization can help estimation and interpretation, it can result in underestimation of the disease burden attributable to the exposure as well as biased comparisons across different exposures and risk factors. Here, we propose sensible PAF estimands for continuous exposures under a potential outcomes framework. In contrast to previous approaches, we incorporate estimation of the minimum risk exposure value (MREV) into our procedures. While for exposures such as tobacco usage, a sensible value of the MREV is known, often it is unknown and needs to be estimated. Second, in the setting that the MREV value is an extreme-value of the exposure lying in the distributional tail, we argue that the natural estimator of PAF may be both statistically biased and highly volatile; instead, we consider a family of modified PAFs which include the natural estimate of PAF as a limit. A graphical comparison of this set of modified PAF for differing risk factors may be a better way to rank risk factors as intervention targets, compared to the standard PAF calculation. Finally, we analyse the bias that may ensue from prior risk factor categorization, examining whether categorization is ever a good idea, and suggest interpretations of categorized-estimands within a causal inference setting.

## 1 Introduction

Population attributable fractions (PAF) are a popular family of metrics in epidemiology for quantifying disease burden attributable to a risk factor at a population level. Most frequently, PAF refers to a binary risk factor, and compares disease prevalence in two populations, one population being observed (perhaps all persons living in the US of at least 18 years of age), with a second hypothetical population that is identical except for the removal of the risk factor of interest. In this regard, PAF attempts to quantify the proportion of current disease prevalence that would be avoided if the risk factor had never been present. The concept was first introduced in Doll and Hill (1952), see Poole (2015) for more details, to estimate the proportion of lung cancer deaths that could be attributed to tobacco-use. Population attributable fractions can vary depending on the source population and on calendar time, primarily according to the prevalence of the risk factor (here tobacco use), but also due to inter-population differences in relative risks. For instance, in the US a recent estimate linking tobacco to cancer (rather than lung-cancer) is 31.7%; however, this PAF is likely to be currently much higher in China, where a higher percentage of individuals smoke, and was higher in the United States in the 1950s when a larger percentage of the population smoked. An important point is that while attributable fractions and related measures such as impact fractions are useful ways to prioritize good risk factor targets for health interventions, they are unfit to measure the benefit of the compared interventions. An obvious reason for this is the implausibility of an intervention completely eradicating the risk factor. The fact that pre-intervention lifetime exposure to certain risk factors can’t be altered may constitute a second more subtle reason. For example, in the unlikely scenario that every American smoker spontaneously decide to abstain from smoking, their cancer-risk subsequent to quitting may still be higher than the corresponding ‘never smokers’ referred to in an attributable fraction.

When risk factors are binary or polytomous, methods to estimate attributable fractions and generate associated confidence intervals are well established for all the main epidemiological study designs including cross-sectional, case-control and longitudinal cohort, see for example Dahlqwist, Zetterqvist, Pawitan, and Sjölander (2016), or Ferguson et al. (2018). However when risk factors are continuous (which we subsequently define as continuous exposures to prevent confusion, with the term risk factor reserved for the discrete case), application and interpretation are not as straightforward as one might at first think. As a result, many researchers resort to dichotomising or categorizing the continuous exposure into a categorical risk factor, resulting in expected underestimation of PAF as we show later in the manuscript. While statistical methods have been proposed to estimate PAF for continuous exposure in the past, our work extends and complements these methods in a number of ways. First, we propose sensible PAF estimands for continuous exposures under a potential outcomes framework. In contrast to previous approaches, we incorporate estimation of the minimum risk exposure value MREV into our procedures. While for exposures such as tobacco usage, a sensible value of the MREV is known, often it is unknown and needs to be estimated. Second, in the setting that the MREV value is an extreme-value of the exposure lying in the distributional tail, we argue that the natural estimator of PAF may be both statistically biased and highly volatile; instead, we propose a family of modified PAFs which include the natural estimate of PAF as a limit. A graphical comparison of this set of modified PAF for differing risk factors may be a better way to rank risk factors as intervention targets, compared to the standard PAF calculation. Finally, we analyse the bias that may ensue from prior risk factor categorization, and suggest interepretations of categorized-estimands within a causal inference setting. To set the scene, we now will review definitions for PAF in the standard, non-continuous framework. In Sections 3 and 4, we will discuss new definitions and estimation methods appropriate for continuous exposures. The manuscript finishes with a discussion in Section 5.

## 2 Methods for binary and categorical risk factors

### Basic definitions

We begin by defining some notation, necessary for subsequent definitions and estimators of attributable fractions. Assume we have collected data on *N* individuals, labeled as *i* = 1, …, *N*. We let *Y*_*i*_ ∈ {0, 1} represent the observed disease outcome for individual, 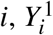, the hypothetical outcome (or potential) assuming individual *i* was exposed to the risk factor, and 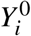, the hypothetical outcome if *i* was unexposed to the risk factor under question. For each individual *i*, we only observe one of the potential outcomes: 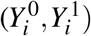, the observed potential outcome being denoted by *Y*_*i*_. While the preceeding statement may seem obvious, it requires technical assumptions that are described in more detail in the Supplementary Material. Under this framework, *P*(*Y* ^0^ = 1) can be directly interpreted as the probability of disease in a hypothetical population with nobody exposed to the risk factor, whereas *P*(*Y* = 1) refers instead to the proportion of the original population with disease.

*PAF* is then defined as:

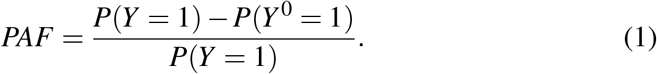

For cohort designs, a slight generalization of (1) has been proposed where two initially disease free populations, one real and one hypothetical population without the risk factor, are followed over time so that attributable fraction is itself a function of time, corresponding to probabilities of the disease event occuring before time *t*, Chen, Hu, and Wang (2006) or Chen, Lin, and Zeng (2010):

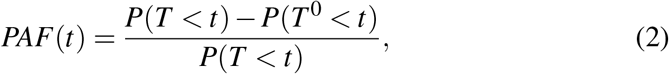

where *T* and *T* ^0^ are observed and counterfactual (assuming absense of the risk factor) survival times. (2) reduces to (1) when fixing *t* and letting *Y* = *I {T < t}*, and *Y* ^0^ = *I {T* ^0^ *< t}*, with *I {A}* representing the indicator function of the event *A*. The difficulty in forming estimators for (1) and (2) lies in estimating *P*(*Y* ^0^ = 1). To proceed, we need to assume the existence of some set of measured covariates *C* ∈ ℝ^*K*^, that when adjusted for suffices to remove confounding between the risk factor, *A*, and response, *Y*. This assumption is often referred to as ‘conditional exchangeability’ and technically implies independence of the potential outcome, *Y* ^0^, and treatment assignment conditional on covariates: *Y* ^0^ ╨ *A*| *C*, see Hernan and Robins (2018) for details. Using the g-formula (again see Hernan and Robins (2018)), it follows that *P*(*Y* ^0^ = 1) = *E*_*C*_(*P*(*Y* = 0 |*A* = 0,*C*)) (where the subscript *E*_*C*_() refers to the expectation over the covariates *C* in the population) so that (1) becomes:

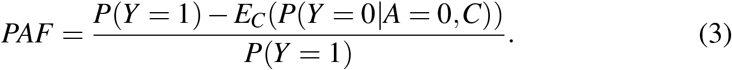

(3) can be used as a basis for the estimation of (1) and (2) in many cross-sectional and cohort sampling designs, provided the sampling of units is representative of the population of interest. The process is to first derive a data-based estimate for the conditional probability of disease given covariates: 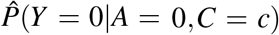, perhaps using logistic regression or survival methods, and then to average this estimate over the empirical distribution of covariates, {*c*_*i*_}_*i*≤*N*_, substituting the result into (1) or (2), Greenland and Drescher (1993) as follows: 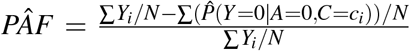. While standardization might constitute the most intuitive approach to estimating *P*(*Y* ^0^ = 1), other approaches such as inverse probability weighting Sjölander (2011), and double robust methods, Sjölander and Vansteelandt (2010), have been suggested in the literature and may be more efficient in some settings.

A clever re-expression of (1), extending the simple formula from Miettinen (1974), was derived in Bruzzi, Green, Byar, Brinton, and Schairer (1985), to facilitate the estimation of PAF in case-control studies. This formula involves averaging the inverse relative risk among the subset of cases that have the risk factor in question according to the following equation:

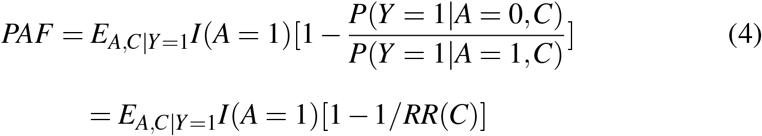

where 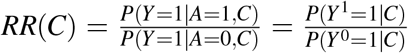 is the relative risk, conditional on *C, I*(*A* = 1) is an 0/1 indicator function taking the value 1 when *A* = 1, and the *E*_*A,C*|*Y*=1_ represents averaging over the distribtuions of *A* and *C* given *Y* = 1. An estimator of *PAF* in case control studies, provided the disease outcome *Y* is rare, can then be derived by substituting a conditional risk-factor/disease odds ratio, 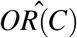 (perhaps estimated from a logistic regression model) into (4), see Benichou and Gail (1990) or Bruzzi et al. (1985) for details. Variance calculations and confidence intervals for estimators based on (3) or (4) can be achieved using the Delta Method, Drescher and Becher (1997), or Bootstrap, Llorca and DelgadoRodríguez (2000). While the above discussion covers the most frequently used modes of attributable fractions, differing methods have been proposed for other study designs, such as population surveys, (Heeringa, Berglund, West, Mellipilán, and Portier (2015)).

### Multilevel exposures

The definitions above can be extended in a straightforward way to multi-category risk factors with at least 3 levels of exposure. As an example, consider a categorization of blood pressure as *high* if both measured systolic BP *>* 140 mmHg and measured diastolic BP *>* 90mmHg, *intermediate* if either systolic BP *>* 140 mmHg or measured diastolic BP *>* 90 mmHg (but not both), and *normal* if both systolic BP ≤140 mmHg and diastolic BP ≤ 90 mmHg. We refer to the level of the risk factor with lowest disease risk as the ‘reference level’, denoting the risk factor levels as 0, 1, …, *L*, with level 0 representing the reference level. Again, we let 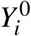 represent the potential outcome supposing individual *i* is unexposed to the risk factor, and 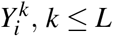, potential outcomes assuming exposure to levels 1, …, *L* of the risk factor. *PAF* can be defined again as (1) and interpreted as the reduction in disease prevalence in a hypothetical population, identical in structure to the population of interest, except that no individual had a non-reference level of the risk factor. Assuming conditional independence of *Y* ^0^ and the assigned level of the risk factor *A* ∈ 0, 1, …, *L* given covariates, the re-expression (3), also holds without alteration. The third re-expression (4), most useful in case-control studies, changes to incorporate relative risks with respect to each of the additional levels of the exposure variable as follows:

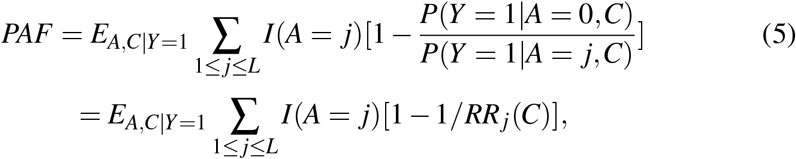

assuming conditional independence of *Y* ^*j*^, *j*≤ *L* and *A*, given *C*. Estimation proceeds as before by substituting estimates for the conditional probability of disease, covariates and exposure into (3) or by instead substituting appropriate estimates of odds ratios or relative risks into (5).

## 3 Extensions to continuous exposure distributions

### Review of some previous work regarding continuous versions of attributable fractions

The Global Burden of Disease (GBD) consortium (see Murray and Lopez (1999)) has carried out a great deal of applied work on estimating attributable fractions and more general measures of disease burden (such as disability adjusted life years) due to a variety of exposures, both discrete and continuous. For continuous exposures, their main strategy is to compare current disease burden with disease burden that might be realized if the distribution of the exposure followed some counterfactual distribution which is considered to be known apriori. This highlights a major difference between their work and the work we propose, where the minimum risk reference level of the exposure is estimated. Differing possible counterfactual distributions are proposed in Murray and Lopez (1999), but the one that most closely corresponds to ‘eliminating the risk factor’ as in (1) is the ‘theoretical minimum’, defined as a counterfactual distribution leading to the lowest probability of disease. In the case of tobacco this might correspond to a population where nobody smoked, but non-deterministic distributions are also possible; for instance, a normal distribution with mean equal to 115 and standard deviation equal to (6) has been proposed for systolic blood pressure, Vander Hoorn, Ezzati, Rodgers, Lopez, and Murray (2004). As Murray and Lopez note, a theoretical minimum distribution is unlikely to be realized in any real population, even through an intervention. More practically realizable distributions are also considered, up to the ‘cost-effective minimum’, a distribution that might be practically achieved through a cost-effective health care intervention. The approach of specifying counterfactual distributions differs from our approach, in that it requires prior expertize regarding healthy levels of the exposure and can make comparison of disease burden due to differing exposures difficult, each needing an individually designed counterfactual distribution. Nevertheless, the consideration of practically realizable interventions is important from a policy maker’s viewpoint. Letting *P*(*x*) and *P*^*^(*x*) represent the actual and theoretical minimum exposure probability distributions, and *RR*(*x*) the relative risk of disease comparing exposure level *x* to the median of *P*^*^ (Note that here *RR*(*x*) is a function of the exposure, unlike equation (4) which described how the relative risk for a binary exposure might interact with covariates), the GBD estimator of attributable fraction is as follows:

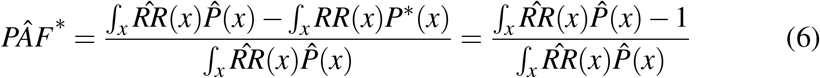

with the second equality assuming that *RR*(*x*) = 1 with probability 1 when *x* is sampled according to *P*^*^. This is essentially the condition that *P*^*^ is truly a risk minimizing counterfactual distribution. *PÂF*^*^ is a continuous extension of Levin’s formula for a binary risk factor, Levin (1952). Unfortuantely if (as is probable) the risk factor disease relationship is confounded, (6) is known to be a asymptotically biased (i.e. inconsistent) estimator of (8) even if adjusted relative risks (conditional on covariates *C*) are substituted for 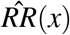, Greenland (1984). This being said, a *PAF* estimator based on (9) or (10) requires estimating the distribution of the distribution of the exposure within cases, direct information for which might be difficult to obtain for rare diseases, especially in lower income countries. The GBD consortium often need to estimate *PAF* based on population level summaries of risk factor exposure, so their reliance on (6) is unsurprising.

An alternative approach to measuring attributable burden due to continuous exposures was considered in Lloyd (1996). Lloyd compared differing values of an exposure, *x*, based on the excess number of disease cases that are observed in comparison to what might be expected if that same group of people (at exposure level *x*) had some baseline value of the exposure, *x*_0_; the excess number of disease cases at exposure value *x* was termed the attributable response. The attributable response can be viewed as a function over *x*, with large values indicating exposure intervals what would especially benefit from an intervention. A decomposition very like that derived in Lloyd (1996), but conditional on confounders, can be found from (10), by setting 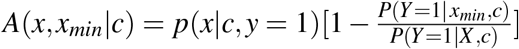, with *p(x* |*c y* = 1) the conditional density of *X* given covariates *C* = *c* and *y* = 1. *A*(*x, x*_*min*_| *c*) is directly proportional to the number of individuals, at covariate level *x* who would be ‘saved’ from disease had they exposure level *x*_0_. *PAF* is then expressed as an integral of this attributable response over the interval of possible exposure values:

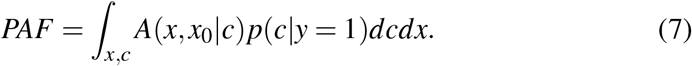

Note that the decomposition used in Lloyd is a little different (and expressed in terms of odds-ratios, rather than relative risks) but the interpretation is the same. In particular, Lloyd doesn’t discuss how to choose the exposure level, *x*_0_, although one of his formulae (the equivalent of (7)) implicitly assumes a monotonic level of risk.

More recently, Traskin, Wang, Ten Have, and Small (2013) describes methods to estimate attributable fractions that incorporate a monotonic relationship between a continuous exposure and disease risk. In contrast to the approaches in Murray and Lopez (1999) and Lloyd (1996) where the estimands are defined directly using conditional expectations, Traskin utliliizes potential outcomes notation to define attributable fractions, and uses constrained logistic regression to enforce monotonic estimated relationships between exposure and the probability of disease. Their work differs from our setting in that it considers a situation where an exposure (measured on a continuous scale) may truly be absent (tobacco usage being an example). In contrast, the setting here considers continuous exposures that may be modified to reduce disease burden but not truly eliminated (such as the effect of blood cholesterol or waist hip ratio on disease risk).

Other work discussing estimators and estimation of attributable fractions in the case of continuously distributed exposures has been rather limited. As an example, Barendregt and Veerman (2010) briefly mention equation (6) as a method to estimate attributable fractions with continuous exposures, although given limited guidance regarding how to employ this formula in practice.

### Definition of *PAF* for a continuous exposure, *X*

For discrete risk factors we defined attributable fractions with reference to a hypothetical population where the risk factor was removed. For continuous exposures, we consider an analogous definition where the distribution of the exposure is fixed at a level which which minimizes disease risk. More formally, consider a continuous exposure, *X* with a population distribution function, *F*(*x*). Let *Y*_*x*_ represent the potential outcome if *X* = *x*, which we assume as before is well-defined (in the Supplementary material we argue that SUTVA assumptions can be more closely satisfied when the exposure is treated as continuous). Consider the function *f* (*x*) = *P*(*Y*_*x*_ = 1). *f* (*x*), assumed continuous in *x*, and can be interpreted as the disease probability if everybody in the population was assigned the exposure *X* = *x*. We also assume the exposure *X* has physiological limits and is as such distributed over a closed interval, implying that *f* (*x*) has a minimum value, *x*_*min*_. This allows us to define *PAF* as:

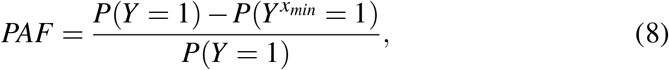

which is a direct generalization of (1). The above definition is usually more appealing when *x*_*min*_ lies well inside the interior of its support. In the case that *x*_*min*_ lies on (or close) to the boundary of the support, interpretation of *PAF* may require consideration of a hypothetical intervention fixing the exposure to an impossible value, and estimation may prove difficult due to paucity of data in the extremes of the *X* -distribution. We will describe methods to address these problems later. Consistent estimation of *PAF* is only possible under similar exchangeability assumptions to the discrete case; that is that we have accurately measured a set of variables *C* satisfying *Y*_*x*_ ╨ *X* conditional on *C*, for all values of *x*. We also note that the minimum risk value, *x*_*min*_, as well as the associated counterfactual probability 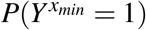 may need to be estimated. The following analogues of (3) and (4) can then be derived:

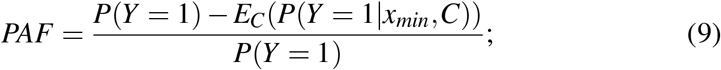

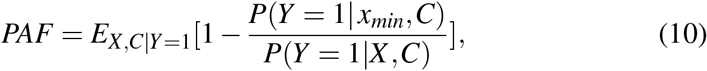

where *P*(*Y* = 1|*x, c*) is defined as the probability distribution of disease given *X* = *x* and *C* = *c*. Provided that estimators of *P*(*Y* = 1|*x*_*min*_,*C*) or 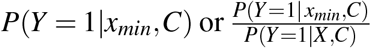 exist (perhaps from a logistic model supplemented with a natural spline for *X*), these estimators can be substituted into (9) or (10) to derive an estimator for *PAF*. Consistent estimation is only possible if the conditional expectation: *P*(*Y* = 1 |*X,C*) is modeled correctly. In the case that *x*_*min*_ is on the boundary of the distribution of *X* estimating *P*(*Y* = 1|*x*_*min*_,*C*) requires extrapolation from this fitted model and the resulting estimator may be both biased and highly variable. Another way to understand this is our data should contain enough individuals having exposure values in a neighbourhood of *x*_*min*_ to stably estimate *P*(*Y* = 1| *X,C*) in that neighbourhood, which is a direct extension of the standard positivity assumptions in causal inference. In general, the optimal value of the exposure, *x*_*min*_, is unknown and needs to be estimated, with obvious estimator being the value *x* achieving minimal estimated risk: *P*(*Y* = ^^^1 | *x,C*) or odds ratio 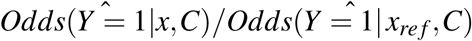 (with *x*_*ref*_ an arbitrary value of the exposure, perhaps the population median). Estimating *x*_*min*_ complicates the construction of a closed form confidence interval for (8), although under certain regularity conditions, most importantly including that *x*_*min*_ is in the interior of the support of *X, PÂF* can be shown to be asymptotically normal with a closed form confidence interval (see the Supplementary material). In this manuscript, standard errors and confidence intervals are instead calculated using Bootstrap methods.

### Comparison to categorized PAF calculations

We now compare the suggested continuous PAF metric above to what is usually calculated when researchers partition the exposure variable into groups. Through such an analysis we can understand under what conditions a grouped PAF statistic is likely to be acceptable. We suppose the support of the exposure, *X*, is partitioned into *K* groups, represented by the factor variable *A* ∈ {0, …, *K* − 1}, with *A* = 0 indicating the group having lowest risk. We assume for simplicity that this group is correctly chosen. For convenience, we define *A*_*k*_ the range of exposure values corresponding to *A* = *k*. When we aprior categorize in this way, before calculating PAF we are in fact estimating:

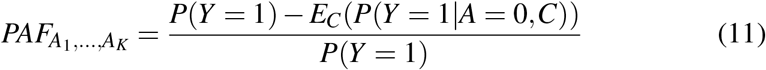

or

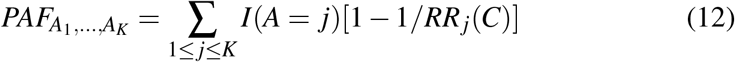

with *RR*_*j*_(*C*) = *P*(*Y* = 1 | *A* = *j,C*)*/P*(*Y* = 1 | *A* = 0,*C*), depending on whether probabilities or relative risks (or Odds Ratios) are easier to estimate. Under the conditional exchangeability assumptions: *Y*_*x*_ ╨*X* |*C*, we show in the Supplementary material that (11) and (12) are equal to:

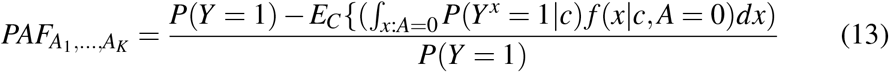

Interestingly, (13) shows that 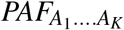 can be interpreted as the proportional decrease in disease prevalence from a randomized intervention where an individual having a vector of confounders *c* is assigned a value of the exposure based on the conditional distribution of *X* given *C* = *c* and *A* = 0.

In the absence of interactions between exposure and covariates, *c* (so that 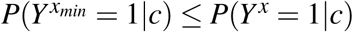) for *x* ≠ *x*_*min*_,

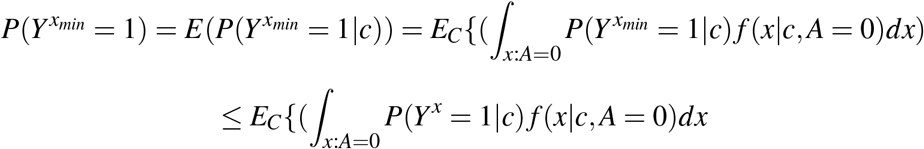

with the result that formula based on (11) and (12) will underestimate *PAF*. The extent of bias is given by the following :

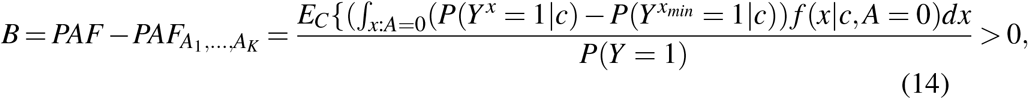

Examining (14) demonstrates that 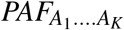 although always smaller than *PAF*, maybe an acceptable proxy provided *P*(*Y*^*x*^ = 1| *c*) is approximately constant in the reference set *A*_0_. In contrast, the larger the variability of disease risk *P*(*Y*^*x*^ = 1|*c*) over *x* ∈ *A*_0_, the greater will be the difference between 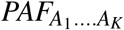 and *PAF*.

### A new family of attributable fraction metrics

Estimating *PAF* as defined by (8) is sensible for risk factors where the exposure disease risk relationship has a well defined minimum in the interior of the exposure’s support. As explained above, estimating *PAF* directly may be difficult when the minimum risk exposure value *x*_*min*_ lies within the extremes of the exposure distribution; indeed in such cases 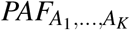 may be a better estimation target. A second alternative that perhaps allows better comparisons of disease burden across risk factors, is to instead consider exposure intervals corresponding to differing lower percentiles of disease risk (somewhat like the couterfactual distributions specified by the Global Burden of Disease team, but specified automatically rather than using biological knowledge) and then predict disease prevalence assuming exposure values were distributed among such a interval. For instance, one might calculate the set of exposure values, *R*_.1_ that corresponds to the lowest 10% of disease risk, in the sense that 10% of the population have exposure values lieing within this range, and the average disease risk at any exposure value outside of this range exceed the average risk for exposure values within the range. We note that estimating the reduction in disease risk if all individuals in the population had exposure values distributed in this interval depends both not just on the range, but also the hypothesized counterfactual distribution within this interval. One convenient choice is to “intervene” only on individuals that originally have exposure values outside the targeted interval, so that in the new hypothetical population they are assigned the closest possible value in the interval to their original value. For instance, if the exposure was wasit hip ratio (as in the data example) and the target interval was (0.799, 0.851), we would assign an individual with WHR 0.9 to 0.851 in the new population. This corresponds to the intervention that minimizes the sum of the absolute value shifts in the exposure values over all individuals within the class of all interventions that successfully move the entire population to the target interval. We define the attributable fraction corresponding to such a counterfactual distribution (for the 100*q*^*th*^ percentile of risk) as *PAF*_*q*_.

More technically, *PAF*_*q*_ has the following definition:

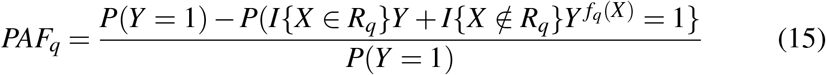

where *R*_*q*_ is the interval of exposure values corresonding to the bottom 100*q*% of risk and *f*_*q*_(*X*) is the closest point in the closure of *R*_*q*_ to *X*. Assuming ignorability (*Y*_*x*_ ╨ *X*|*C*) for all *x* in the support of the exposure, and the assumption that the potential outcomes, 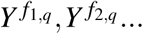 at any boundary points: *f*_1,*q*_, *f*_2,*q*_, … of *R*_*q*_ are equidistributed, conditional on any *C* = *c, PAF*_*q*_ can be reexpressed or

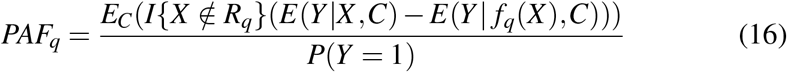

and

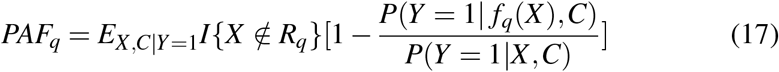

(see the Supplementary appendix for details). To estimate *PAF*_*q*_, one can substitute logistic regression based estimates of *P*(*Y* = 1| *f*_*q*_(*X*),*C*) or *P*(*Y* = 1| *f*_*q*_(*X*),*C*)*/P*(*Y* = 1 | *X,C*) into (16) or (17) for various values of *q*, and then average over the empirical distribution of covariates, *C*. Note that as *q* decreases to 0, *PÂF*_*q*_ converges to *PÂF* and similarly, *PAF*_*q*_ converges to *PAF* (see the supplementary material), but the estimators will become more variable. Why would one bother with this, when one can estimate PAF directly? There are two main reasons why this family of PAF-metrics may be useful. First, over most *q* ∈ (0, 1) *PAF*_*q*_ may be better estimated than *PAF* (*PÂF*_*q*_ will tend to have a lower variance and bias particularly when *x*_*min*_ is in the extremes of the *X* -distribution. Second, for reasonable values of *q* (perhaps values of *q >* 0.1, although there is some subjectivity as to what constitutes ‘reasonable’), *PÂF*_*q*_ may correspond more closely to the degree of risk elimination that a real-world intervention of the risk factor might achieve. For these reasons, we argue that plots based on estimated *PÂF*_*q*_ (with the quantile *q* on the x-axis) may be a more useful way to compare risk factor disease burden compared to simply estimating *PÂF*. An example of such a plot is included in the data-example shown in Section 4.

## 4 Practical Application

### Three continuous exposures from the INTERSTROKE dataset

We illustrate the ideas discussed in the previous section with examples regarding the burden of stroke due to naturally continuous exposures: waist hip ratio (WHR), measured Alternative Healthy Eating Index diet score (AHEI diet score) and ApoB/ApoA ratio, based on INTERSTROKE, a large international case control study, O’Donnell et al. (2010). Our analysis includes data for the 13,462 cases and 13,483 controls collected during the period from March 2017 until May 2018. O’Donnell et al. (2016) described PAFs using (5), according to a division of the exposure values into risk-factor tertiles. Their implicit model included adjustments for age, gender and region (through conditional logistic regression) as well as smoking status, regular physical activity, Diabetes Mellitus, alcohol intake, psychosocial stress factors, and the presence of cardiac risk factors. The models considered here are similar, but instead model waist hip ratio, diet score and ApoB/ApoA with cubic splines. We also consider slightly differing sets of covariates for the models representing each risk factor. In assessing the effect of diet score, our adjustment set consisted of age, geographic region, sex, stress, physical activity, smoking status and alcohol, while when assessing the effects of ApoB/ApoA and waist hip ratio, the adjustment set includes the adjustment set for diet score in addition to diet score, hypertension, waist hip ratio and ApoB/ApoA. These differing adjustment sets represent an attempt to exclude variables that are potentially downstream effects of the exposure of interest from the adjustment set, although in practice results varied only slightly as a result of these differences. The spline models (adjusted as described) are displayed graphically below, and are natural cubic splines with 3 inner knots (at the 25th, 50th and 75th percentiles) for all three exposures and outer knots at the 0.1th and 99.9th percentiles for Waist Hip Ratio and Diet Score, with the upper outer knot for ApoB/ApoA at the 95th percentile (a biologically implausible sharp decline in the estimated OR was observed when the upper knot was put at the 99th percentile). The relationships between ApoB/ApoA and diet and stroke risk appear monotonic. In contrast, there is a hint that stroke risk might increase at very low values of waist hip ratio, although there is no obvious biological reason why that might happen.

### Using *PAF*_*q*_ to graphically compare INTERSTROKE exposures

The splines in Figure 1 display estimated adjusted Odds Ratios for stroke comparing various exposure levels to the respective population medians: 0.93 for waist hip ratio, 0.78 for ApoB/ApoA and 22.19 for diet score. The x-axis interval on each plot spans from the 1st to the 99th percentile of the exposure. The relationships for diet score and ApoB/ApoA appear monotonic, apart from a untrustworthy kink in the ApoB/ApoA curve at high values of the exposure, and as a result *x*_*min*_, the minimum risk value of exposure, is in the extreme of the distribution. As explained in Section 3, in this situation Odds Ratios (or probabilities) comparing risk at *x*_*min*_ and other possible exposure values have high variability (as is apparent from the widening of the confidence intervals in Figure 1 at the extremes), which propagates into unstable estimation of *PAF*, (8), and confuses the comparison among different exposures. Here we instead demonstrate the estimation of *PAF*_*q*_. Recall, the intervals 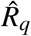 associated with *PÂF*_*q*_ over a set of percentiles *q*, correspond to the lowest 100*q*% of disease risk. These are demonstrated for *q* ∈ 10%, 30% and 50% in Figure 2. For instance, for waist hip ratio, the intervals corresponding to the lowest 10%, 30% and 50% of disease risk are (0.799,0.851), (0.749, 0.893) and (0.709, 0.931) respectively.

**Figure 1:**
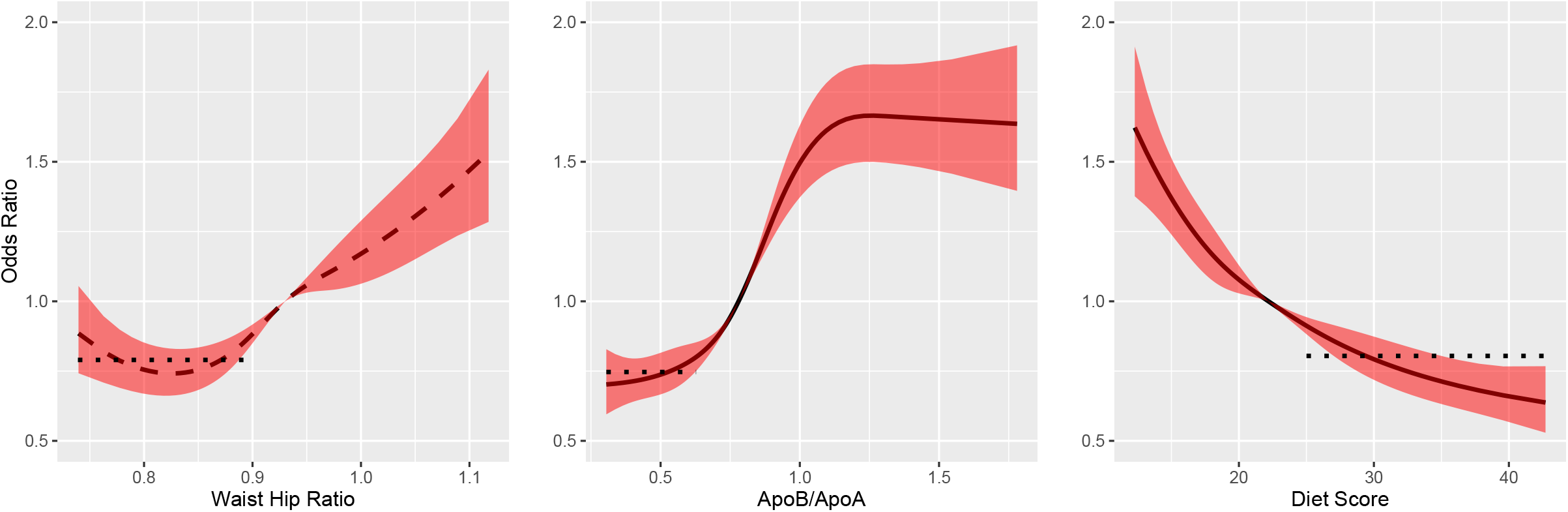
Natural cubic splines and pointwise 95% confidence bands showing adjusted odds ratios (compared to median exposure) for waist hip ratio, ApoB/ApoA and diet score. The dotted line represents the average odds ratio among cases in the first tertile of Waist Hip Ratio and ApoB/ApoB and in the top tertile of diet score.

**Figure 2:**
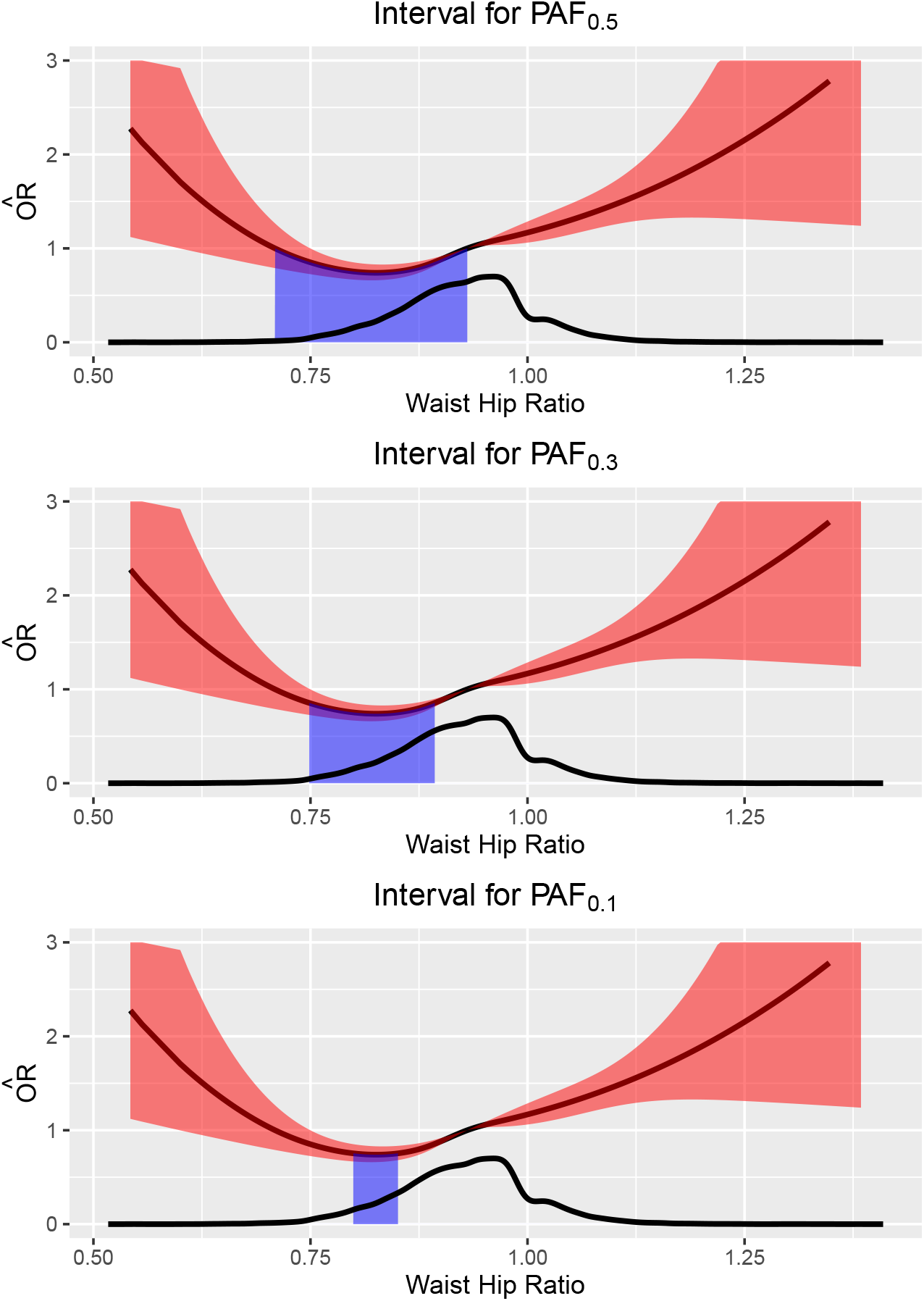
Intervals corresonding to *PAF*_0.1_,*PAF*_0.3_ and *PAF*_0.5_ for waist hip ratio.

The corresponding estimates of *PAF*_*q*_ for a range of *q* between 0 and 1 are shown in Figure 3. Here, while diet score has the highest estimated *PAF* (as evidenced from Figure 3 and Table 1 for small *q*) it is a poor estimate with a wide confidence interval, as we might expect when *x*_*min*_ is extreme (see Table 1). In contrast, a comparison of *PAF*_*q*_ for the three exposures (see Figure 3) indicates that, concerns about risk factor modifiability aside, ApoB/ApoA may be a better intervention target, despite its estimated PAF being lower than that of diet score. The respective shapes of the three curves is also of interest. An asymptotic plateau of *PÂF*_*q*_ as *q* → 0 indicates an exposure where the minimal risk region is broad and well defined (such as waist-hip ratio or ApoB/ApoA), and the asymptote of the curve, that is the *PAF*, can be estimated. In contrast, *PÂF*_*q*_ rapidly increases as *q* → 0 for diet score, indicating that the minimum-risk is not well defined and estimation of *PAF* maybe unstable.

**Table 1:** *PÂF*_*q*_ for AHEI diet score, Waist Hip Ratio and ApoB/ApoA calculated using INTERSTROKE. 95% confidence intervals (calculated with Bootstrap) are in parentheses. Note that attributable fractions are displayed as percentages rather than proportions in this table.

| q | AHEI diet score<br>(%) | Waist Hip Ratio<br>(%) | ApoB/ApoA<br>(%) |
| --- | --- | --- | --- |
| 0.5 | 8.07 (5.01, 10.74) | 6.57 (3.76, 9.46) | 16.56 (14.31, 19.03) |
| 0.3 | 13.89 (11.00, 17.11) | 14.92 (11.75, 18.24) | 22.91 (20.05, 25.86) |
| 0.1 | 24.64 (19.08, 29.51) | 22.77 (17.64, 28.41) | 27.51 (23.04, 31.43) |
| 0.05 | 29.79 (24.15, 36.09) | 23.74 (18.78, 29.39) | 28.69 (23.75, 33.53) |
| 0.01 | 37.62 (28.15, 47.39) | 24.09 (18.51, 29.76) | 29.77 (21.10, 38.13) |
| 0.001 | 43.67 (25.09, 61.23) | 24.11 (18.77, 29.97) | 30.31 (19.63, 42.71) |

**Figure 3:**
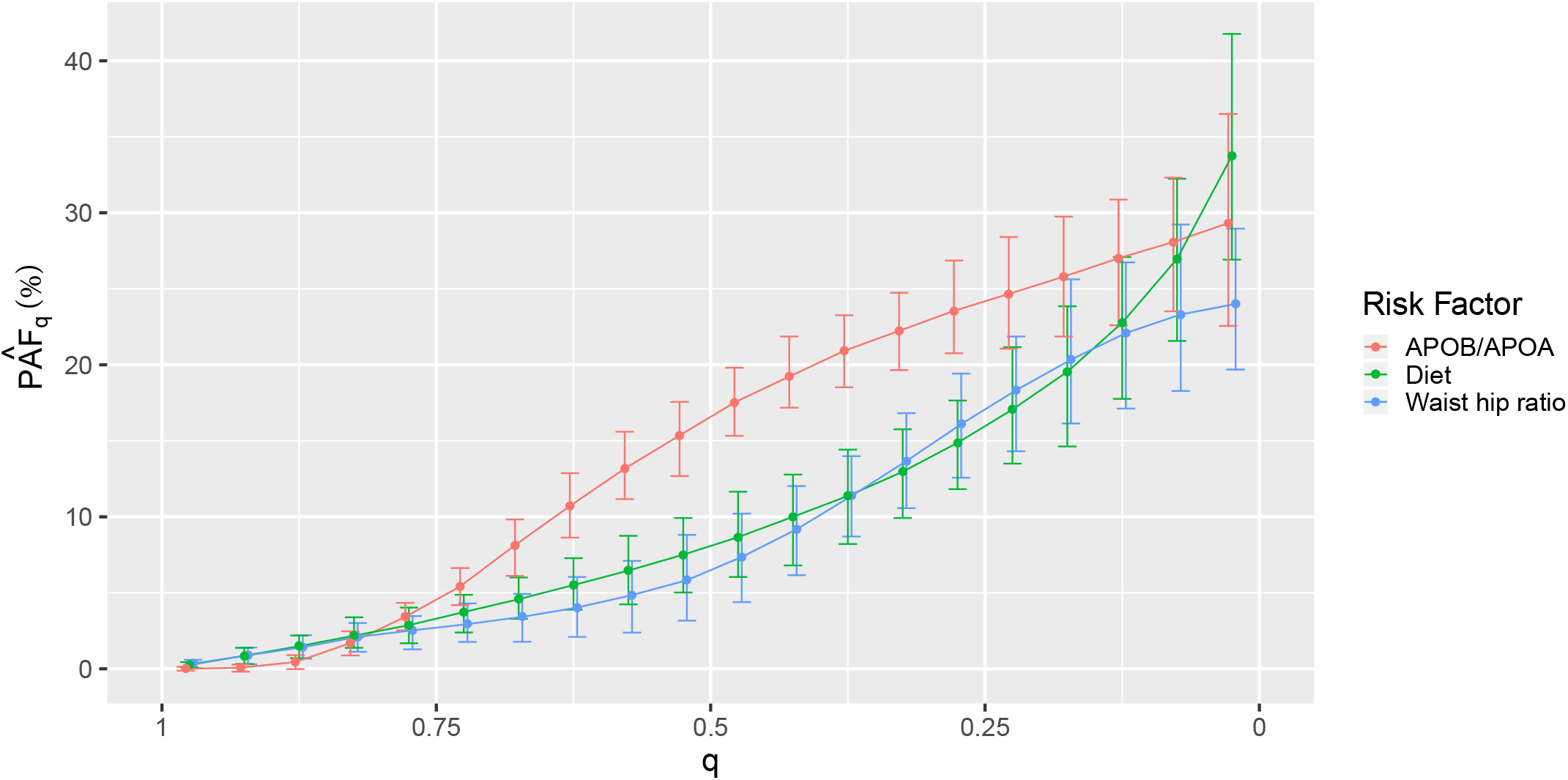
Estimates of *PAF*_*q*_ (as a percentage rather than a proportion) for ApoB/ApoA, Diet Score and Waist hip ratio. 95% Confidence intervals were calculated via Bootstrap. Notice that as *q* → 0, *PAF*_*q*_ → *PAF*.

### Previous categorical analysis

In the original INTERSTROKE analysis (O’Donnell et al. (2016)), diet score, ApoB/ApoA and waist hip ratio were compared using tertiles. Table 2 compares this calculation to *PÂF*_.001_ (which in turn should be a good approximation of (8)). We write the estimated PAF with a tertile grouping as 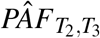. As expected from the analysis in Section 3, an approximation using tertiles is always smaller than *PÂF*_.001_ and the two are most discrepant for diet score. Some insight regarding these calculations can be gleaned from Figure 1 where the estimated average OR (compared to median exposure) in the lowest risk tertile within cases is represented as a dotted line. As explained in Section 3, in situations where a calculation with tertiles gives an acceptable approximation of *PÂF*, the difference between this dotted line and the minimal value of the OR (that is the OR comparing the minimum risk exposure to the median exxpousre) should be relatively small. This discrepancy is larger for diet score than for waist hip ratio or ApoB/ApoA ratio which explains the extent to which *PÂF*_.001_ and 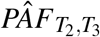 differ for diet score.

**Table 2:** 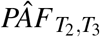 is the result of applying formula (13) to the three INTER-STROKE exposures, divided into tertiles, with estimated odds ratios replacing relative risks. *PÂF*_0.001_ is an estimate of (15) with *q* = 0.001. Note that attributable fractions are displayed as percentages rather than proportions in this table.

| Exposure | $\hat{PAF}_{0.001}(\%)$ | $\hat{PAF}_{T_2, T_3}(\%)$ |
| --- | --- | --- |
| AHEI diet score | 43.67 (25.09, 61.23) | 21.1 (15.4-26.9) |
| Waist Hip Ratio | 24.11 (18.77, 29.97) | 19.1 (13.7-24.5) |
| ApoB/ApoA | 30.31 (19.63, 42.71) | 27.2 (21.5-32.9) |

## 5 Summary and Discussion

While estimating attributable fractions for continuous exposure distributions is not a new problem, we hope to have provided a novel perspective in this paper. Our methods differ from previous approaches (for instance Murray and Lopez (1999)), in that in our approach optimal exposure values (or ranges) are estimated by relationships in the data, in contrast to being pre-defined counterfactual distributions.

The analysis we describe highlights a dichotomy between exposures where the relationship with disease risk is monotonic, and those where the relationship with disease is U-shaped. When the relationship is U-shaped, there is a well defined minimum risk exposure value and in some sense PAF is well defined. Alternatively, when the relationship is monotonic, the optimum setting for exposure may be in the extremes of the distribution, and estimating risk ratios using such an extreme value as a reference may suffer from both extrapolation and high variance, which filters into the estimated attributable fraction: *PÂF*. In particular, *PÂF* may give misleading comparisons for disease burden when comparing multiple risk-factors. When comparing exposures that have a monotonic relationship with disease risk, graphically examining the slightly altered metric, *PÂF*_*q*_ as a function as *q* ranges from 1 to 0 may give a better indication of exposures that are likely to benefit from an intervention.

An important point that arises from our analysis is that the bias due to prior categorization of an exposure into groups depends on the extent that disease risk varies among exposure values in the reference group. This degree of bias will vary over differing categorized exposures even if the groups are defined in similar ways for each exposure (for instance, using tertiles or other quantiles of each exposure), indicating that comparing *PAF* may be flawed even in these situations. On the other hand, PAF comparisons across categorized risk factors may be reasonable if disease risk conditioned on differing values of each exposure shows limited variation across the chosen reference groups even when this reference group is chosen differently across different exposures.

Admittedly, the estimation methods described here could be improved. The approach described here to estimate *PAF* substitutes regression based estimators of *P*(*Y* = 1|*x*_*min*_, *c*) or 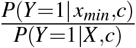, averaged over the empirical covariate distribution into (9) or (10), and calculates standard errors via Bootstrapping. However, an estimating equation approach involving both an outcome probability model, *P*(*Y* = 1 *x*, | *c*), and model for exposure assignment given covariates, *f* (*x*| *c*) could in theory be derived. The solutions to such estimating equations can under certain conditions be shown to be doubly robust, that is asymptotically consistent if either the outcome model or the exposure model is correct, and in addition may in some situations be more efficient even when both models are correct. In addition, the sandwich formula suggests the asymptotic standard error for such an estimator negating the need to use the Booststrap. Such an approach would extend the work of Sjölander and Vansteelandt (2010), who derived doubly robust estimates of the regular attributable fraction in cohort and case control datasets. While such an estimating approach in the continuous exposure case treated here, unless *x*_*min*_ is known we would not expect the approach to be double robust, as bias within *P*(*Y* = 1|*X,C*) would lead to mis-estimation of *x*_*min*_.

This work was supported by a grant from the Health Research Board of Ireland [EIA-2017-017]

## Data Availability

R code for performing analyses described in the manuscript can be obtained by contacting the corresponding author

