## Supplementary material for "Population attributable fraction for continuously distributed exposures"

March 4, 2020

#### 1 SUTVA assumptions for PAF

To unambiguously define $Y_i^0$ in formula (1), Stable Unit Treated Value (SUTVA) assumptions of treatment variation irrelevance and no interference are made, see [1] for details. These assumptions may often be questionable in situations where attributable fractions are calculated. For instance, consider the example of stroke burden attributable to waist hip ratio as described in the main manuscript. Differing potential outcomes may result if high waist hip ratio originates from genetic, dietary or reasons based on lifestyle, indicating that $Y^0$ may not be unambiguously defined. Similarly, regarding no-interference, perhaps an individual's risk of lung cancer is lessened if her partner gives up tobacco. In these scenarios, PAF can still be imagined and estimated as referring to a hypothetical population where nobody was obese or nobody smoked, but the calculated PAF would refer to the versions of the risk factor (and lack of risk factor) in the collected data, and not necessarily to any real world intervention on the risk factor. While we recognize these problems and ambiguities, we adopt a potential outcome framework in the manuscript as despite these caveats it provides a more coherent basis for attributable fraction definitions. In addition, regarding SUTVA assumptions, the consideration and development of methods that respect the undelying continuity of the exposure of interest are certainly a step in the right direction. For instance, suppose an underlying exposure $X$, having well defined potential outcomes $Y^x$ is categorized into $A \in \{0,1\}$ depending on whether or not $X \geq 0$. Then, while $Y^x$ is well defined, $Y^{A=0}$ and $Y^{A=1}$ are not. The interpretation of the resulting attributable fraction (calculated according to the values of $A$) is the percentage prevalence reduction from a randomized intervention that assigns a random value of the exposure $X$ according to the conditional distributions within $Y=1$ and $A=1$.

### 2 Confidence intervals

While we have used Bootstrap techniques to find confidence intervals for $PAF$ in the examples in the main manuscript, when the minimal risk exposure level, $x_{min}$, is well defined and can be estimated as the unique stationary point of the function $f(x) = P(Y^x = 1)$, it is possible to derive analytic confidence intervals using the theory of M-estimators (see [2] for instance). M-estimators solve for the zero of a multivariate function relating the data and true parameters, constructed so that the population value of the function (or equivalently expectation of the function) is 0. For instance, suppose the relationship between the response, $Y$, covariates, $C$, and exposure $X$ is modeled with a logistic regression. Parameter estimates can be found by solving the $p$ equations defined by setting the sample score vector $\sum_{i\leq N} S_i(\beta) = \sum_{i\leq N} \frac{\delta}{\delta\beta} log f(Y_i|x_i, c_i, \beta)$ to 0. In addition, $x_{min}$ needs to satisfy the equation: $\sum_{i\leq N} \frac{d}{dx} P(Y = 1|c_i, x_{min}) = 0$, or equivalently $\sum_{i\leq N} \frac{d}{dx} logOdds\{P(Y=1|c_i, x_{min})\} = 0$. Sometimes, the assumption of no interactions between exposure and covariates will be made. Then the portion of the linear predictor that depends on the exposure is typically modelled via a basis expansion $\sum_{j\leq p} \gamma_j B_j(x)$, with $\gamma$ being a subvector of $\beta$ and this second equation simplifies to: $\sum_{j\leq p} \frac{d}{dx} \gamma_j B_j(x_{min}) = 0$, the dependence on

$i$ now removed. Finally, we need an equation (or equations) that relates the parameters $(\beta, x_{min})$ to $PAF$, which we represent here by the parameter $\pi$. This will typically vary depending on the study design. For case control designs, the appropriate equation is $\sum_{i\leq N} I(Y_i = 1)(1 - \frac{P(Y_i=1|x_{min},c_i)}{P(Y_i=1|x_i,c_i)}) - \pi = 0$. For prospective designs, 3 equations might instead by used; namely $\frac{1}{N}\sum_{i\leq N} Y_i - \pi_1 = 0$, $\frac{1}{N}\sum_{i\leq N} P(Y = 1|x_{min}, c_i) - \pi_2 = 0$ and $\pi - \frac{\pi_1 - \pi_2}{\pi_1} = 0$. In either case, we relabel this set of equations as the vector equation $\sum m(Y_i, X_i, C_i, \beta, x_{min}, \pi) = 0$ or $\sum m(Y_i, X_i, C_i, \beta, x_{min}, \pi_1, \pi_2, \pi_3) = 0$, with different components of $m$ corresponding to the subequations defined by the score vector, minimum risk level $x_{min}$ and $\pi$. Solving the system will lead to the estimates of $PAF$ suggested in this manuscript. Under reasonably mild conditions detailed in [2], it follows that the joint parameter estimator $(\hat{\beta}, \hat{x}_{min}, \hat{\pi})$ is asymptotically normal with mean $(\beta, x_{min}, \pi)$ and variance $E(\frac{d}{d\theta}m(Y_i, X_i, C_i, \theta))^{-1}E(m(Y_i, X_i, C_i, \theta)m(Y_i, X_i, C_i, \theta)^T)E(\frac{d}{d\theta}m(Y_i, X_i, C_i, \beta, \theta))^{-1}$ with $\theta$ representing the entire parameter vector. Data based estimates of the components of this variance matrix, and hence the $PAF$ can be estimated via the Sandwich estimator.

### 3 Derivation of estimation formula for $PAF_q$

Here we show that the counterfactual formula for $PAF_q$ that is:

$$PAF_q = \frac{P(Y = 1) - P(I\{X \in R_q\}Y + I\{X \notin R_q\}Y^{f_q(X)} = 1\}}{P(Y = 1)}$$

can be re-expressed as:

$$PAF_q = \frac{E_C(I\{X \notin R_q\}(E(Y|X, C) - E(Y|\, f_q(X), C)))}{P(Y = 1)}$$

under the assumptions of exchangeability $Y^{f_{j,q}} \perp\!\!\!\perp X|C$ for boundary points $f_{j,q}$ $j = 1, ...,$ of $R_q$ with $f_q(X)$ defined as the closest boundary point to $X$. We also assume the boundary points of $R_q$ have equi-distributed counterfactuals, that is: $Y^{f_{j,q}}|C = c \sim Y^{f_{1,q}}|C = c$, for all boundary points $f_{j,q}$.

#### Proof:

First note that $Y^{f_q(X)} = \sum_j I(f_q(X) = f_{j,q})Y^{f_{j,q}}$. It follows that

$$\begin{aligned}
&Y^{f_q(X)}|X = x, C = c \sim \\
&\sum_j I(f_q(X) = f_{j,q})Y^{f_{j,q}}|X = x, C = c \sim \\
&\sum_j I(f_q(X) = f_{j,q})Y^{f_{j,q}}|C = c \sim \\
&Y^{f_{1,q}}|C = c
\end{aligned}$$

for all $x$ and $c$. Where the second last statement is from the above conditional exchangeability and the last statement from the equi-distributed counterfacturals.

Now, let $B(X) = I(X \in R_q)$. Note that the numerator of $PAF_q$ involves

$$\begin{aligned}
&P(B(X)Y + (1 - B(X))Y^{f_q(X)} = 1) = \\
&E_{C,B}(P(B(X)Y + (1 - B(X))Y^{f_q(X)} = 1|C, B(X)\}) = \\
&E_{B(X),C}(P(B(X)Y + (1 - B(X))Y^{f_q(X)} = 1|C, B(X)\}) = \\
&E_C(P(B(X) = 1|C)P(Y = 1|C, B(X) = 1) + P(B(X) = 0|C)P(Y^{f_q(X)} = 1|C, B(X) = 0))
\end{aligned}$$

Note also that:

$$P(Y^{f_q(X)} = 1|C, B(X) = 0) =$$
$$E_{X|B(X)=0}P(Y^{f_q(X)} = 1|C, X) =$$
$$E_{X|B(X)=0}P(Y^{f_{1,q}} = 1|C = c) =$$
$$P(Y^{f_{1,q}} = 1|C = c) =$$
$$P(Y^{f_{1,q}} = 1|C = c, X = f_{1,q}) =$$
$$P(Y = 1|C = c, X = f_{1,q})$$

again by conditional exchangeability, so that the above becomes:

$$P(B(X)Y + (1 - B(X))Y^{f_q(X)} = 1\} =$$
$$E_C(P(B(X) = 1|C)P(Y = 1|B(X) = 1, C) + P(B(X) = 0|C)P(Y = 1|C = c, X = f_{1,q})$$

Now writing:

$$P(Y = 1) = E_C(P(B = 0|C)P(Y = 1|B = 0, C)+P(B = 1|C)P(Y = 1|B = 1, C),$$

and writing $PAF_q = \frac{P(Y=1)-P(B(X)Y+(1-B(X))Y^{f_q(X)}=1\}}{P(Y=1)}$, the result follows by simple algebra. The proof for equation (13) in the main manuscript follows along similar lines.

### 4 $PAF_q \rightarrow PAF$ as $q \rightarrow 0$, assuming that $X$ is a continuous random variable and $f(x)$ is a continuous function

(Here we assume the technical condition that $R_q$, the set of risk factor values of measure equal to $q$ with the smallest supremal value of $f$, is uniquely defined for each $q > 0$. This will be the case if the set $y_x = \{x : f(x) = y\}$is finite for each $y$), or informally that the graph $f(x)$ is 'nowhere flat'.

First note that by the definition of $PAF$ and $PAF_q$, the statement follows directly if we can prove $P(Y^{f_q(X)} = 1) \downarrow P(Y^{x_{min}} = 1)$ as $q \downarrow 0$. Suppose this is not the case; then for some $\epsilon > 0$, $P(Y^{f_q(X)} = 1) \geq P(Y^{x_{min}} = 1) + \epsilon$, for all $q > 0$.

Next, since $f(x)$ is continuous at $x_{min}$, we can choose $\delta$ be such that if $|x - x_{min}| < \delta$, then $|f(x) - f(x_{min})| < \epsilon$

Now, since $X$ is continuous (more technically absolutely continuous w.r.t. Lebegue measure), we have that

$$P(S_\delta) = P\{|X - x_{min}| < \delta\} = q^* > 0.$$

By definition $R_q \subset R_{q^*}$ if $q < q^*$(recall $R_q$ is the set of risk factor values of measure equal to $q$ with the smallest maximal value of $f(x)$). Supposing then that $q' < q^*$, $S_\delta = \{x : |x - x_{min}| < \delta\}$ is a set of $P$-measure larger than $R_{q'}$ and as a result we can choose $x^* \in S_\delta / R_{q'}$. Again by the definition of $R_{q'}$, $f(x^*) \geq f(x)$ for all $x$ in $R_{q'}$. It follows that $f(x^*) \geq P(Y^{f_{q'}(X)} = 1)$, since $P(Y^{f_{q'}(X)} = 1)$ can be calculated as a weighted average of values of $f(x)$, with $x$ in $R_{q'}$. But $f(x^*) < f(x_{min}) + \epsilon$ by the continuity of $f$. This in turn implies that $P(Y^{f_{q'}(X)} = 1) < P(Y^{x_{min}} = 1) + \epsilon$, a contradiction.

### 5 Proof of formula for discretized estimation

Here we prove the equality of equations (14) and (15) in the main manuscript, under conditional exchangeability. That is:

$$\frac{P(Y = 1) - E_C(P(Y = 1|A = 0, C = c))}{P(Y = 1)} \tag{1}$$

$$= \frac{P(Y=1) - E_C\{(\int_{x:A=0} P(Y^x=1|c)f(x|c,A=0)dx)}{P(Y=1)}$$

when $Y_x \perp\!\!\!\perp X|C$ for all $x$. Note that:

$$\begin{aligned}
E_C(P(Y &= 1|A=0,C)) \\
&= E_C E_{X|A=0,C}((P(Y=1|A=0,C=c,X)) \\
&= E_C \int_{x:A=0} P(Y=1|A=0,C=c,X=x)f(x|c)/P(A=0|c) \\
&= E_C \int_{x:A=0} P(Y^x=1|A=0,C=c,X=x)f(x|c)/P(A=0|c) \\
&= E_C \int_{x:A=0} P(Y^x=1|C=c)f(x|c)/P(A=0|c)
\end{aligned}$$

where the 4th equality uses consistency and the 5th conditional exchangeability. Inserting the above into (1) gives and noting $f(x|c)/P(A=0|c) = f(x|c,A=0)$ gives the result.

### References


[1] Hernan and Robins. *Causal Inference.* Boca Raton: Chapman & Hall/CRC, Forthcoming, 2018.

[2] Anastasios Tsiatis. *Semiparametric theory and missing data.* Springer Science & Business Media, 2007.
